# Modelling Palliative and End of Life resource requirements during COVID-19: implications for quality care

**DOI:** 10.1101/2020.07.23.20160564

**Authors:** Daniel Chalk, Sara Robbins, Rohan Kandasamy, Kate Rush, Ajay Aggarwal, Richard Sullivan, Charlotte Chamberlain

## Abstract

**Background:** There were between 84,891 and 113,139 all-cause excess deaths in the United States (US) from February 1^st^ to 25^th^ May 2020. These deaths are widely attributed directly and indirectly to the COVID-19 pandemic. This surge in death necessitates a matched health system response to relieve serious health related suffering at the end of life (EoL) and achieve a dignified death, through timely and appropriate expertise, medication and equipment. Identifying the human and material resource needed relies on modelling resource and understanding anticipated surges in demand.

**Methods:** A Discrete Event Simulation model designed in collaboration with health service funders, health providers, clinicians and modellers in the South West of England was created to estimate the resources required during the COVID-19 pandemic to care for deaths from COVID-19 in the community for a geographical area of nearly 1 million people. While our analysis focused on the UK setting, the model is flexible to changes in demand and setting.

**Results:** The model predicts that a mean of 11.97 hours (0.18 hours Standard Error (SE), up to a max of 28 hours) of additional community nurse time, up to 33 hours of care assistant time (mean 9.17 hours, 0.23 hours SE), and up to 30 hours additional care from care assistant night-sits (mean of 5.74 hours per day, 0.22 hours SE) will be required *per day* as a result of out of hospital COVID-19 deaths. Specialist palliative care demand is predicted to increase up to 19 hours per day (mean of 9.32 hours per day, 0.12 hours SE). An additional 286 anticipatory medicine bundles or ‘just in case’ prescriptions per month will be necessary to alleviate physical symptoms at the EoL care for patients with COVID-19: an average additional 10.21 bundles (0.06 SE) of anticipatory medication per day. An average additional 9.35 syringe pumps (0.11 SE) could be needed to be in use per day (between 1 and 20 syringe pumps).

**Conclusion:** Modelling provides essential data to prepare, plan and deliver a palliative care pandemic response tailored to local work patterns and resource. The analysis for a large region in the South West of England shows the significant additional physical and human resource required to relieve suffering at the EoL as part of a pandemic response.

**Why Was This Study Done?:** The resource required for the relief of suffering at the EoL in the community setting has been poorly described. The stark mortality resulting from the COVID-19 pandemic has highlighted the essential requirement to better understand the demand and available supply of EoL resource to prepare, plan and deliver a palliative care pandemic response.

**What Did the Researchers Do and Find?:** This manuscript describes the first open access model to describe EoL resource need during COVID-19 and presents an analysis based on a UK population of nearly 1 million people. The model identified a large increase in need for staff time, including registered community nurses, health care assistants and specialist palliative care nurses and doctors, as well as pressure on resources including syringe pumps and anticipatory medication (such as opioids) used at the EoL for symptom relief from breathlessness and delirium.

**What Do These Findings Mean?:** The model findings are critical in planning for a second wave of COVID-19. The open-access nature of the model allows researchers to tailor their analysis to low and middle income or high-income settings worldwide. The model ensures that EoL care is not an afterthought in pandemic planning, but an opportunity to ensure that the relief of suffering at the EoL is available to all.

## Introduction

465,740 people have died since the 30^th^ of January 2020 when the WHO Director General declared COVID-19 a Public Health Emergency of International Concern.(1) 41.5% (193,366) of COVID-19 deaths to date have been in Europe,(1) but the burden in low and middle income settings is increasing exponentially. Increasingly, excess mortality is being used to describe the impact of COVID-19, comparing current mortality with deaths at the same period in previous years, indicating deaths that are likely related to COVID-19, but that have not been directly attributed to it due, primarily, to differences in testing. Excess mortality also includes deaths due indirectly to COVID-19, resulting from disruptions to health services and health seeking because of the pandemic. Excess mortality in the United States(US) from February 1^st^ 2020 to 25^th^ May estimates between 84,891 and 113,139 all-cause excess deaths.(2) Cumulative excess mortality in the United Kingdom (UK) is estimated to be around 48,823 (31^st^ January to week ending 15^th^ May).(3) In the peak week of COVID-19 deaths in April in the UK, there was a 113% higher excess mortality compared with the same period in the previous 5 years.(4) This surge in death rates is substantial and represents an extensive personal and societal cost, as well as a significant increased burden on end of life care resource. It is essential that those dying from COVID-19, as with all illness, receive high quality relief of suffering, which is mandated by the human right to health and a recognised component of essential universal health coverage.(5) Without access to the essential palliative care package, countries will not meet the sustainable development goals, and inequity of access to quality End of Life (EoL) care is likely to increase.(5, 6)

In COVID-19, where predominant physical symptoms at the EoL include delirium and respiratory distress(7) palliative care has a key role in administration of anticipatory medicines. Anticipatory medication may include midazolam for agitation, morphine or equivalent opioid for breathlessness or pain, amongst others. Palliative and EoL care are critical in the pandemic context to support the psychological and spiritual needs of patients, their loved ones and health professionals particularly in the context of social isolation due to the virus. EoL care, defined in this paper as the last days of life, differs by culture, country and by setting (home, hospital, hospice, care home). Lower income countries frequently rely more on informal care networks of family and friends who in some care settings are supported by health professionals with an interest in palliative and EoL care and may not be able to access essential palliative medicines.(5) High income countries rely on a specialist and generalist model of delivery of palliative care. In the UK, generalist palliative care, provided by General Practioners (GP’s) and Community nurses (also known as District Nurses) working in peoples’ residences and care homes support EoL care for the majority of the population.(8) Specialist palliative care provision varies across the UK, but includes hospice services, hospital palliative care teams and community palliative care. The critical resource components of EoL care in any setting include multidisciplinary EoL expertise (for instance physiotherapy, occupational therapy, doctors, nurses, health care assistants, spiritual care, social work support and psychologists), as well as availability and access to anticipatory medication, and the delivery systems of those medication (e.g. commonly syringe pumps in a UK setting). Despite increasing recognition of the importance of palliative care during the pandemic,(9-11) identifying the essential resource to continue the high quality, equitable delivery of such care has been overlooked in many national responses.

Mathematical modelling allows estimation of required resource in the face of changing demand. It is common in infectious disease and acknowledged for its utility in planning and coordination for predicted surges in demand for healthcare resource, changing epidemiology of the disease and social responses to it. The model presented here was conceived to predict local EoL resource demand for the anticipated waves of COVID-19 death. It was developed in collaboration with clinicians, local health service funders (known as commissioners in the UK) and health providers and based on representative UK health data for a population of 968,314 in the South West of England.(12) It predicts key end of life care resources needed for a given level of incoming activity (dying) and duration of EoL care, adaptable to different geographical regions within the UK or other countries, and flexible, allowing users to include newly emerging evidence about COVID-19. The model is free and open source and has been made publicly available.

A palliative care pandemic response requires proactive planning around four elements: ‘staff, stuff, space and systems’.(13) Delivering this palliative pandemic framework requires essential estimation of demand for end of life resources in each of these elements - this can be facilitated by EoL resource modelling and is necessary to facilitate equitable access to the high quality relief of suffering essential in a pandemic.

## Methods

A conceptual model was designed in collaboration with practicing clinicians, modelling experts and local health service funders and providers, including links with the Clinical Cabinet who represent local senior leaders in clinical decision making. This collaborative effort aimed to estimate essential EoL care resource need (approximated in ‘bundles’ of EoL resource) in the different settings in which it is delivered. This information was then used to develop a Discrete Event Simulation model. Discrete Event Simulation models entities flowing between discrete, sequential events, and the resource utilisation needed to meet incoming levels of activity.(14) Probability distributions are used to emulate the real world variability observed in arrival and process times. The model, using the Python-based SimPy (15) framework, is available here: https://colab.research.google.com/drive/1K0-xsqG_uIpoWhw86tjOUM55OcFfhq_2.

Figure 1 shows an overview of the basic model structure. Patients enter the model at the point they need EoL care at home, in a care home or in hospital. The time between patients arriving into the model in each setting is randomly selected from an exponential probability distribution and can be set to increase or decrease over time to account for growth or decline in activity. An exponential distribution is commonly used to model time between independent events occurring, such as the arrival of people into a system. The model allows for the capture of non-COVID-19 patients, but we only simulated COVID-19 patient activity to predict excess resource need. We excluded two resources from our analysis – Hospital Nurse time (which is already incorporated in hospital planning) and GP time (as we were unable to quantify excess GP time for EoL care as a result of COVID-19). Both are included in the model should others wish to specifically include them in their analyses. The important spiritual and psychosocial contributions (social workers, chaplains) of the palliative care MDT were not included in the model due to limited or absent data for modelling.

**Figure 1.**
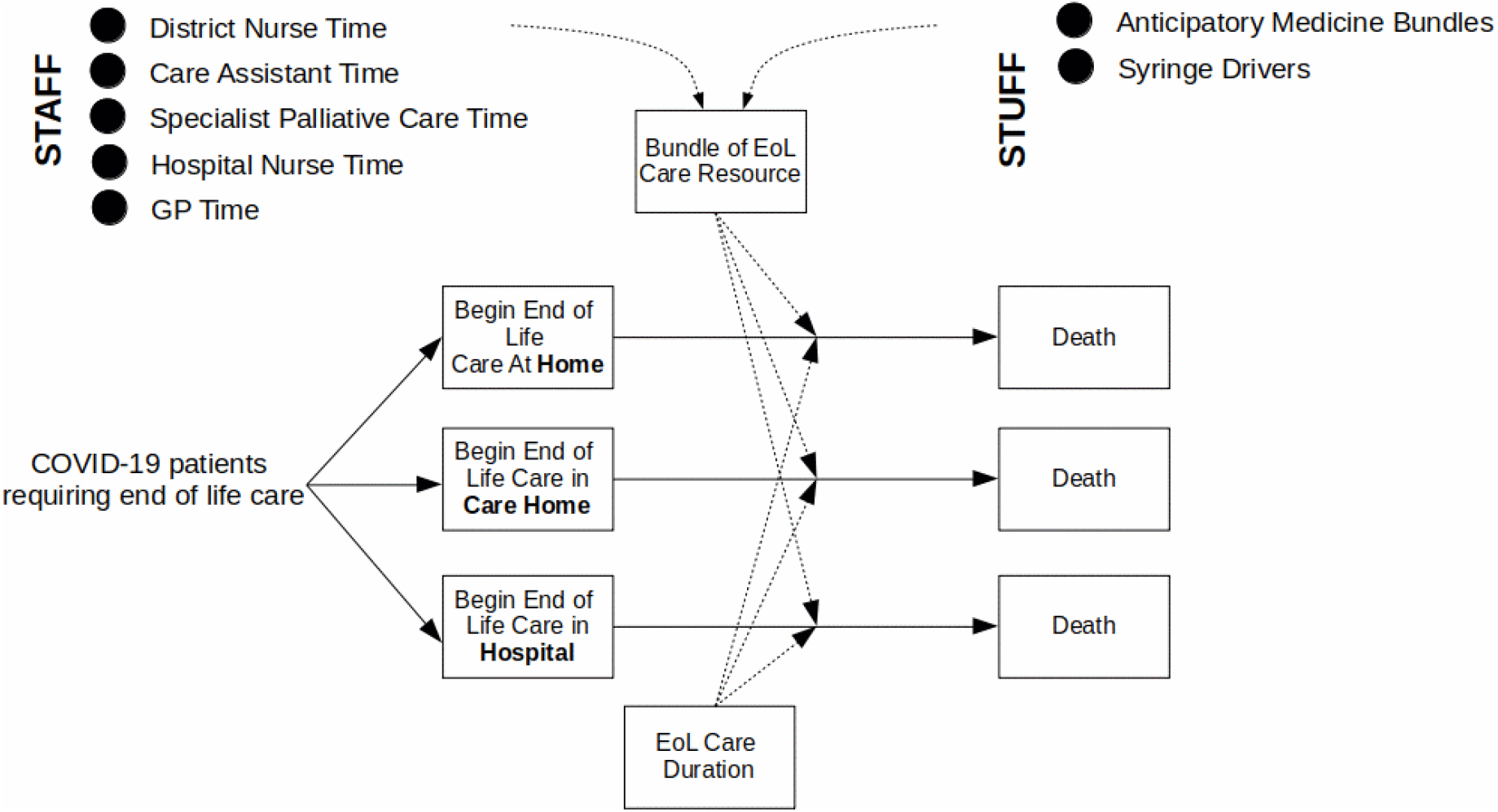
Overview of the basic structure of the model.

Patients are allocated a “bundle” of EoL care resources, each of which has a probability of being selected, and a duration for their EoL care, sampled from an exponential distribution. There are seven types of resource in the model: anticipatory medicine bundles, syringe pumps, community nurse time, hospital nurse time, health care assistant time for personal care, specialist palliative care time (for example local hospice support) and GP time. Anticipatory medicine bundles may include a combination of midazolam, morphine sulphate and an anti-emetic, but these have deliberately not been specified since the bundle may vary per region and the significance is that the drugs are generally prescribed together for EoL care and therefore they have been treated as a bundle. Staff resources are specified in terms of the number of “visits” (physical or virtual) per day, the length of visits, and the resources needed per visit. Visit lengths are sampled randomly for each patient from a normal distribution, in which most values are assumed to be close to the mean. The necessary resources needed over time are recorded. The simulation can be run for a chosen length of time, and for a chosen number of runs. This is important when there is randomness in the model, to ensure that results aren’t solely based on runs of “good” or “bad” luck in random number selection. The model calculates results over runs to maximise accuracy.

To model the EoL care resource needs for Bristol, North Somerset and South Gloucestershire, we derived parameter values from a variety of local and national sources. Office for National Statistics data(16) indicates that, from 31^st^ March 2020 to 3^rd^ April 2020, the total COVID-19 deaths recorded was 35, with 29 recorded in hospital. Government statistics(17) report 130 new cases in the locality over the same four days. Therefore, we estimate the percentage of patients requiring EoL care in hospital is 22.3% (29 / 130).

Data from Scotland(18) indicates 24.6% of COVID-19 deaths take place in care homes and 13.3% at home. This would estimate 46.8 new cases per day in the locality, with 11.5 per day in care homes and 6.2 per day at home. Assuming that 22.3% of cases require EoL care, then we would estimate 6.47 new EoL care cases per day in hospital, 2.56 per day in care homes, and 1.38 per day at home. This translates to mean times between arrivals of new patients in the model of 223 minutes for hospital cases, 563 minutes for care home cases and 1,043 minutes for cases at home. We modelled a stable mean rate of activity based on the “peak” levels of activity observed at the time of writing.

A study of 101 COVID-19 patients in London found that patients spent an average of 2 days under the palliative care team.(7) The local palliative care team estimated the community EoL care duration to be similar, at around 2 days for those in care homes, and 3 days for those at home.

Table 1 shows an overview of the bundles of resource used in the model. For COVID-19 patients receiving EoL care at home, it was estimated that around 25% of patients would need the highest level of care, due to limited function or multiple complex needs. They would typically receive two one-hour district nurse contacts per day, and we assume a standard deviation of 15 minutes around each contact time. Two care assistants visiting for 40 minutes (standard deviation 5 minutes) four times per day are needed for personal care, as well as one care assistant on a 12 hour “night sit” every other day. Around 1 hour of Specialist Palliative Care per day would be needed, along with a bundle of anticipatory medicines and a syringe pump to administer medicines.

**Table 1.** Overview of the bundles of resource used in the model for each of the three care settings, including their frequencies, and the amount of each resource required.

| Setting and magnitude of resource need (high, medium and low) | Frequency Within Care Setting | District Nurse | Care Assistant (CA) | Night-Sit Care Assistant | Specialist Palliative Care | Anticipatory Medicine Bundles | Syringe Pumps |
| --- | --- | --- | --- | --- | --- | --- | --- |
| **HOME High EoL Care Need** | 25% | 2 visits per day x 1 hour (15 minute SD) | 2 CA x 4 visits per day x 40 minutes (5 minute SD) | 1 CA x 12 hour night sit every other day | 1 hour per day (15 minutes SD) | 1 for duration | 1 for duration |
| **HOME Medium EoL Care Need** | 70% | 1 visit per day x 30 minutes (5 minute SD) | 1 CA x 3 visits per day x 30 minutes (5 minute SD) |  | 1 hour every other day (15 minutes SD) | 1 for duration | 1 for duration for 50% of patients |
| **HOME Low EoL care Need** | 5% | 1 visit every other day x 30 minutes (5 minute SD) |  |  |  | 1 for duration |  |
| **CARE HOME (Residential Home, External Support Required)** | 17.3% | 2 visits per day x 1 hour (15 minute SD) |  |  | 1 hour per day (15 minutes SD) | 1 for duration | 1 for duration |
| **CARE HOME (Nursing Home, External Support Required)** | 22.7% |  |  |  | 1 hour per day (15 minutes SD) | 1 for duration | 1 for duration |
| **CARE HOME (Residential or Nursing, Low EoL Care Need)** | 60% | 1 visit every other day x 30 minutes (5 minute SD) |  |  |  | 1 for duration |  |
| **HOSPITAL** | 100% |  |  |  |  | 1 for duration | 1 for duration |

Around 5% of patients at home have very low-level needs, or do not want regular input. For these patients, anticipatory medicine bundles would still be provided, and a minimal level of District Nurse contact, estimated at around 30 minutes every other day (standard deviation 5 minutes). The remaining 70% at home would receive one 30 minute District Nurse contact per day (standard deviation 5 minutes). Three 30 minute (standard deviation 5 minutes) care assistant visits per day, and a bundle of anticipatory medicines would be provided. Around half of the patients in this cohort require a syringe pump. Specialist Palliative Care support was estimated at around 1 hour (standard deviation 15 minutes) every other day. Travel time was excluded from visit duration for estimates of community nurses, care assistants and specialist palliative care support, as travel times can vary widely and travel time is already considered as part of community care planning.(19)

For patients in care homes, those in residential homes would typically require the highest level of EoL care resource because of the lack of internal nursing support. Therefore, we used our three bundles of high, medium and low-level resource for care home patients to represent those in residential homes needing external support, those in nursing homes needing external support, and those in either nursing or residential homes who have very low level needs, respectively. In addition, we would expect to see more patients needing EoL care in nursing homes due to the complexity of their care needs. A non-weighted mean calculation from 2011 Census data shows an approximate 50/50 split between nursing and residential home residency in the locality.(20) A prior study reported 41,969 annual deaths in nursing homes, compared to 32,138 deaths in residential homes.(21) This represents a 31% higher death rate in nursing homes, assuming population levels are the same. The CCG estimated around 40% of care home residents require non-trivial external EoL care support. If the nursing home cohort is 31% larger, we estimate that 17.3% of patients fit into the “residential home – external support required” bundle, and 22.7% into the “nursing home – external support required” bundle.

For those in a residential home needing external support, two 1-hour district nurse contacts and 1-hour specialist palliative care support (15 minutes standard deviation) would be required per day. These patients would also be supplied with a bundle of anticipatory medicines and a syringe pump. Patients in a nursing home would only need district nurse support to answer short adhoc queries over the phone, so we exclude them from this bundle. Patients with low level needs would only require a bundle of anticipatory medicines, and minimal district nurse contact, estimated at around 30 minutes (standard deviation of 5 minutes) every other day. Patients receiving EoL care in hospital would receive their nursing and personal care support via the hospital staff, and so the only external resources required would be syringe pumps and bundles of anticipatory medicines.

We ran the model for a simulated 28 days in each model run. The model was allowed to “warm up” for 5 days in each run first to account for the model starting from an empty state, and so results from the first 5 days in each run are excluded. The model was run 100 times, and average results taken across the batch of runs.

## Results

### Staff

The model predicts that up to 28 hours of additional district nurse time (mean of 11.97 hours, 0.18 hours Standard Error (SE)) will be required per day because of out of hospital COVID-19 deaths. 95% of days are predicted to need no more than 19 hours of additional district nurse time and 5% of days would need less than 5.7 hours. Up to 33 hours of care assistant time would be required per day (mean 9.17 hours, 0.23 hours SE), with 95% of days needing no more than 19 hours but only 5% days needing less than 2 hours. In addition, up to 30 hours additional care from night-sits per 24 hour period may be required (mean of 5.74 hours per day, 0.22 hours SE), with 95% of 24 hour periods requiring no more than 17 hours, and around 26% of nights needing no night-sit resource.

Specialist palliative care demand is predicted to increase up to 19 hours per day (mean of 9.32 hours per day, 0.12 hours SE). 95% of days are predicted to need no more than 14 hours, but only 5% of days would need less than 5 hours.

### Stuff

Physical resource demand will increase by an additional 286 anticipatory medicine bundles or ‘just in case’ prescriptions per month because of demand for EoL care for patients with COVID-19 both in the community and in hospital. This represents an average additional 10.21 bundles (0.06 SE) of anticipatory medication per day, with a predicted minimum and maximum of 2 and 23 bundles per day, respectively. 95% of days are predicted to need no more than 16 bundles, but only 5% of days would need less than 5 bundles.

For syringe pumps, an average additional 9.35 pumps (0.11 SE) would be needed to be in use per day, with a predicted minimum and maximum of 1 and 20 pumps in use per day, respectively. 95% of days are predicted to need no more than 15 pumps in use, but only 5% of days would need less than 5 pumps in use. Figures 2 and 3 show the model’s predicted mean resource usage per day for each of 28 days in the model for ‘staff’ and ‘stuff’ resources, respectively, averaged across the 100 runs in the trial. Error bars show standard errors of the mean across the runs in the trial.

**Figure 2.**
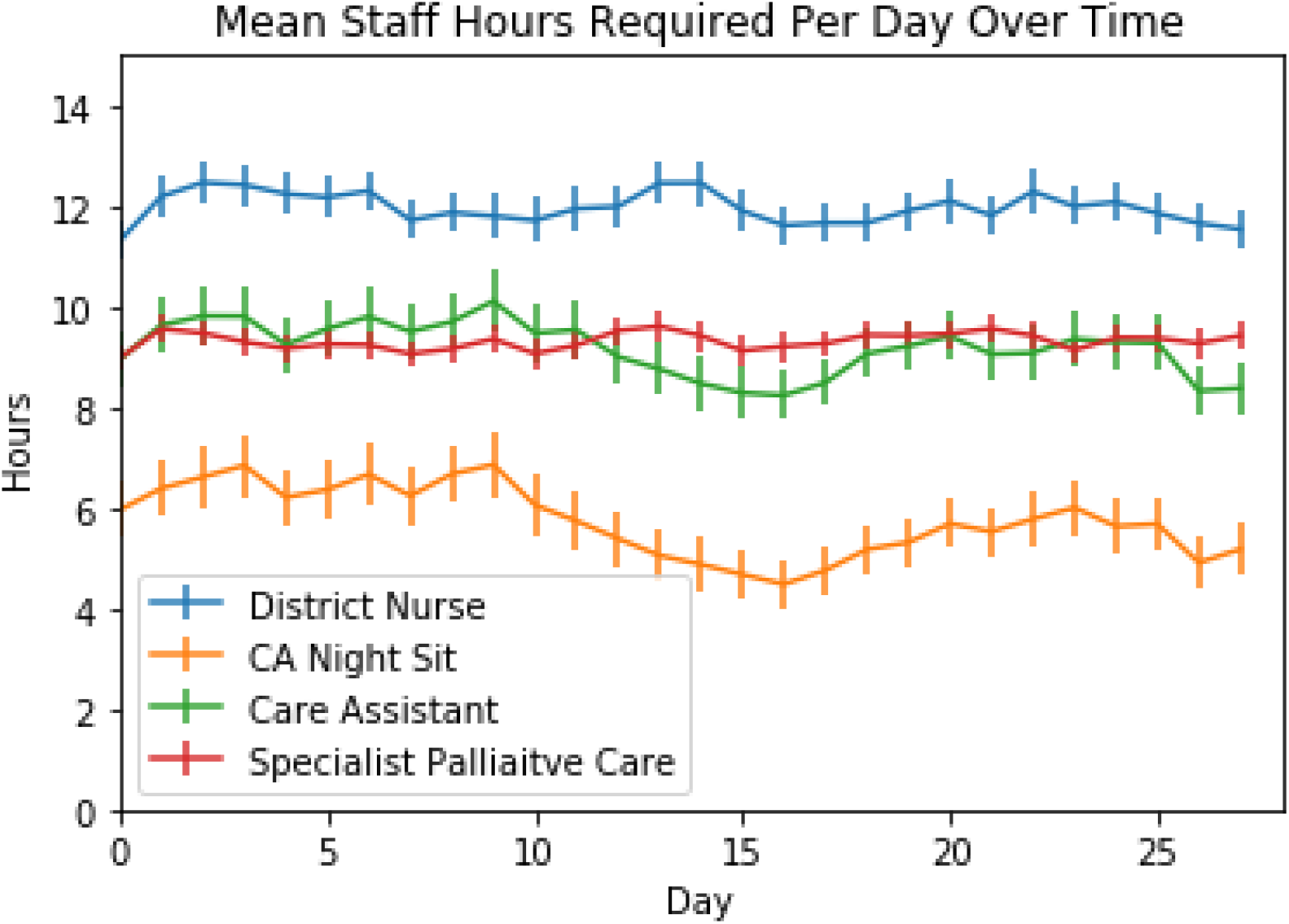
Predicted mean staff hours required per day across the 100 runs of 28 days of the simulation for community nurses, night sits, care assistants and specialist palliative care.

**Figure 3.**
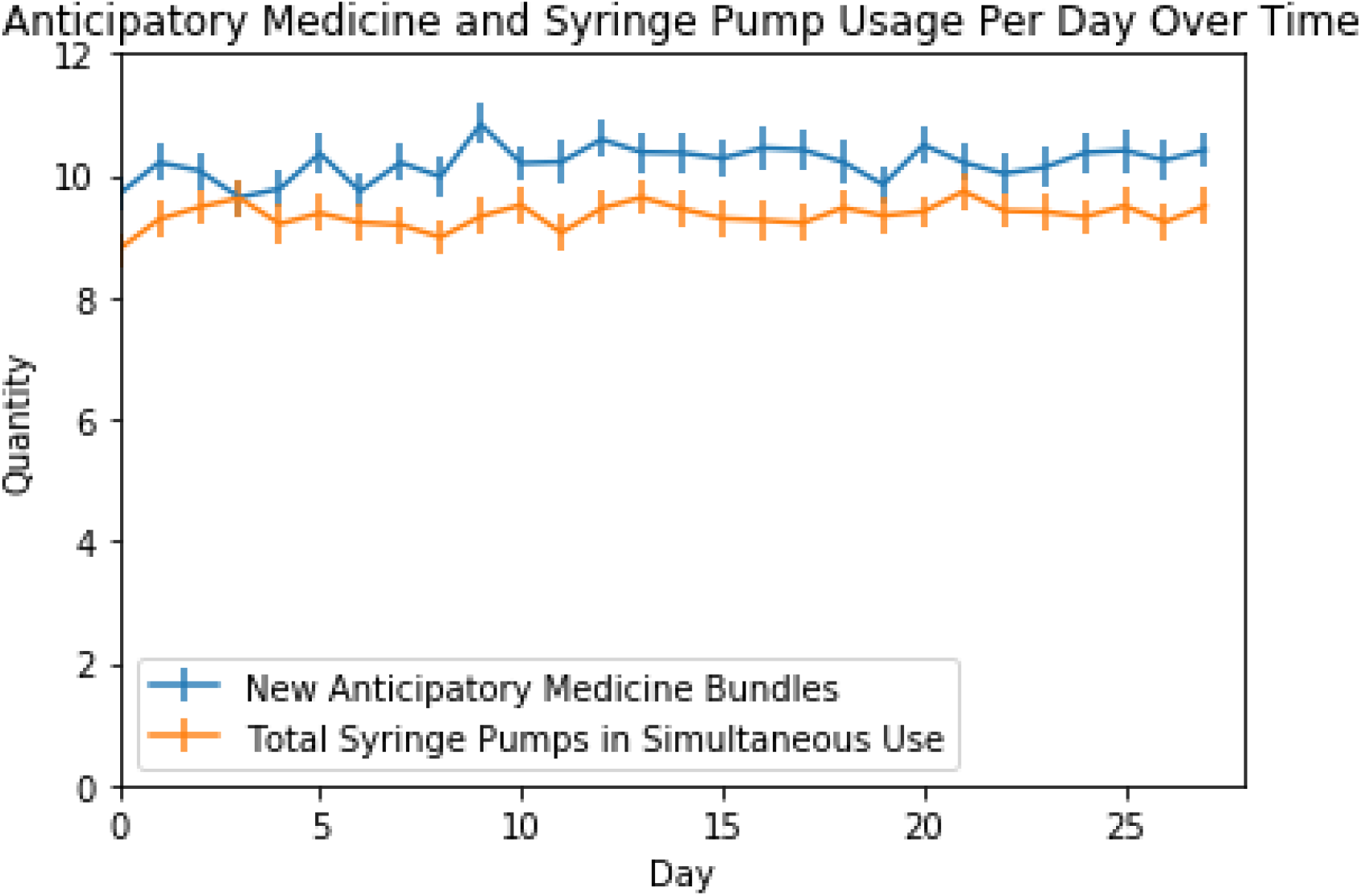
Predicted mean anticipatory medicine and syringe pump usage per day across the 100 runs of 28 days of the simulation. Anticipatory medicine numbers represent numbers of new bundles required per day, whilst syringe pump numbers represent total needed to be in simultaneous use per day.

## Discussion

### Main findings

An increase in community nurse, care assistant, specialist palliative care time and anticipatory medicine and syringe driver resource is predictable with the rise in deaths due to COVID-19. The significance of this model is in quantifying that rise across a population of nearly 1 million in the UK to demonstrate the importance of planning and preparation for EoL care resource need during a pandemic. With an average of 12 hours extra district nurse time and nine hours of additional care assistant time *per day*, care clearly cannot be met with current staffing levels, let alone with anticipated staff absence due to sickness, household isolation, other caring responsibilities and with a rise in non-COVID-19 deaths as has been demonstrated worldwide.(2, 4)

Based on the model analysis for additional COVID-19 EoL care need, specialist palliative care demand is predicted to increase by around 9 hours per day. This will necessitate different ways of working. 286 additional anticipatory medicine bundles or ‘just in case’ prescriptions predicted to be needed per month, due to COVID-19 EoL care, which in the UK context cannot be reused even if they are not used by the named patient, may risk specific drug shortages. Nine additional syringe pumps needing to be in use per day, at a cost of around £1742 each or £15,678 in total is also a significant added resource demand for EoL care for patients with COVID-19 in the community. Nine syringe pumps are also only the average in use in a 24 hour period and to ensure demand is met 95% of the time 15 would be required, at a total cost of £26,130.

### Comparison with the literature

To our knowledge this is the first published model of EoL resource need during the COVID-19 pandemic worldwide. Understanding demand for resource to relieve serious health related suffering at the EoL is critical in providing essential solace to patients in knowing they are not abandoned, with care provided up to death and beyond for their families. Palliative care is important to mitigate the moral distress in health professionals who may be in the unenviable position of triaging patients to palliative care support where limited intensive care support may be available.(22) Palliative care is consistently shown to reduce symptom burden at the EoL and improve bereavement outcomes for families.(23, 24) Planning for the predicted surge in demand follows the Downar and Seccareccia pandemic plan(13) identifying ‘stuff’, such as ensuring sufficient anticipatory medication and syringe pumps (and subcutaneous butterflies) availability, and acknowledging the minimum specialist and generalist palliative care staff required to fufill our duty of care, alleviating serious health related suffering. In many countries, like the UK, EoL community data are challenging to obtain because the community setting crosses different health and social care partnerships, as well as third sector charitable organisations. It is hardly surprising, therefore, that the clear increased risk in terms of acquisition and survival from COVID-19 to those who live in care homes was recognized late, and appropriate allocation of resources was hindered in the UK.(25)

Community driven carer injectable drug administration policies,(26) intended to provide symptom relief and access to medication even where there may be staff shortages is one solution introduced in this locality to address the predicted staff shortages. Rapid generation of national and local guidelines on topics such as advance care planning, communication in COVID-19, EoL symptom care and bereavement support can and have been delivered remotely using digital solutions, demonstrating the large scale collaborative efforts through national and international organisations(27, 28) that are possible to upskill other health professionals and mitigate the capacity constraint of specialist palliative care. Anticipating the large increases in demand in anticipatory medication and syringe pumps has also contributed to innovative community solutions, such as altered prescribing behaviour to prevent drug shortages and a greater reliance on butterfly needles to administer subcutaneous injections.

### Strengths and limitations

This model is a step change in planning for EoL care during the COVID-19 pandemic and provides detail for the stuff, staff, space and systems pandemic planning approach. It has been made freely available to enable modellers and health service funders to estimate need for EoL resource in different regions, countries and for different rates of disease and resource estimations for usual care. The model also highlights the gaps and opportunities for research in EoL care in the community. The work was undertaken collaboratively with health service funders, providers, clinicians and modellers to base assumptions in the reality of current practice during the pandemic.

Some of the limitations of the model assumptions are limitations due to available data, rather than of the model per se. The rate of COVID-19 death in the community setting is based on the latest available figures at the time of the project, and where those for England were absent, the most geographically similar area available: Scotland, was inputted. While all deaths from COVID-19 are a tragedy, it has been postulated that between 5% and 15% of COVID-19 deaths may have occurred in people who would have died of other causes within the year.(29) However, even if this was the case and that all deaths from COVID-19 do not represent *additional* resource and only 85% extra is required, the increased demand of COVID-19 deaths has occurred acutely, rather than spread out over the longer time period of a year, and therefore the supply constraint is still very problematic. The model does not take into account excess mortality which has not been attributed directly to COVID-19. In other words, those deaths that have erroneously not been identified as COVID-19 due to absent testing, or deaths that have taken place because usual care has been disrupted leading to delayed healthcare presentation, or interrupted local management of illness. When comparing the observed deaths in the UK for March and April with the expected number of deaths for the same period, based on the average deaths usually occurring in that time (over a 5 year period) it is clear that deaths due to COVID-19 are significantly increased, but deaths which are not being directly attributed to COVID-19 have also risen significantly.(3) While many of the non-COVID deaths may in reality be unconfirmed COVID-19 deaths, the flexibility of the model will allow more accurate resource estimations to provide for palliative and EoL care to be made with improved data over time and this will become the focus of further work.

While we would have liked to have included the resource requirement for EoL care in the community for non-COVID-19 deaths and to compare current observed practice with expected practice based on historic evaluation, the resource data were too limited and in the first instance we have focused on the deaths from COVID-19. Similarly, inclusion of GP time is felt to be of high importance, but data on time spent by GPs on EoL care is limited and worthy of further research given the significant role they play in the care of those at the EoL in the community. Hospice inpatient services have also not been included in this analysis (although the model could easily accommodate their inclusion). This is because hospice was treated as a finite resource, which was not affected by the pandemic in that there are limited numbers of inpatient hospice beds. For instance, in our locality there are 25 inpatient beds and only one patient has been confirmed to have died with COVID-19 in a hospice setting. Increased community support via redeployment of hospice health professionals to telephone advice lines was accounted for in the analysis as a specialist palliative care resource. Personal protective equipment (PPE) has not been included in the model. Potential COVID-19 changes to the anticipatory care bundle, such as more carer administered drugs (e.g. lay carer administration of injectable medication or increased prescribing of non-injectable medication to mitigate staffing shortages) was not included in the analysis in order to model the care which most clearly mirrors that which is deemed optimal practice. Finally, we have not included “travel time” for community nurses and carers, therefore in more rural settings this will change estimates, which the model can accommodate.

Models can be retrospectively validated and adapted once data of the impact matures.

### Implications for practice

Our cultural bias towards health care heroics risks overlooking the essential contribution of palliative and EoL care to a pandemic. Armed with the structure to outline palliative care pandemic planning,(13) this study allows realistic modelling of the essential ingredients to prepare, plan and deliver a palliative care pandemic response tailored to local work patterns and resource. Without anticipating the resource constraints, equitable care is compromised. This pandemic is a reminder of the vital need for collaborative, flexible working and quality data collection to inform preparation and planning to prevent deaths with physical and psychosocial distress. Emphasising EoL care does not negate the importance of life-saving or even life-sustaining care, but acknowledges the moral imperative to provide care for everyone in a pandemic, even where a cure is not possible.

## Data Availability

All data used is described in the paper, and the model described in the paper has been made available free and open source (link provided in the manuscript)

https://colab.research.google.com/drive/1K0-xsqG_uIpoWhw86tjOUM55OcFfhq_2

## Acknowledgements

DC is funded by the National Institute for Health Research Applied Research Collaboration South West Peninsula and CC supported by the National Institute for Health Research. This report is independent research funded by the National Institute for Health Research Applied Research Collaboration South West Peninsula. The views expressed in this publication are those of the author(s) and not necessarily those of the National Institute for Health Research or the Department of Health and Social Care. We would like to acknowledge the support of Dr Alexander Middleditch for supporting questions around COVID-19 hospital data for local acute trusts

